# Hydroxychloroquine pre-exposure prophylaxis to prevent SARS-CoV-2 among health care workers at risk for SARS-CoV-2 exposure: A nonrandomized controlled trial

**DOI:** 10.1101/2022.07.01.22277058

**Authors:** Vanessa N. Raabe, Andrew Fleming, Marie I. Samanovic, Lilin Lai, Hayley M. Belli, Mark J. Mulligan, H. Michael Belmont

**Author notes:** Corresponding Author: Vanessa Raabe, 430 E 29^th^ St., Alexandria Center for Life Sciences West Tower, 3^rd^ Floor, New York, NY 10016.

## Abstract

**Background:** Aerosol-generating procedures increase the risk of severe acute respiratory syndrome coronavirus 2 (SARS-CoV-2) infection among health care workers (HCWs). An effective pre-exposure prophylaxis would mitigate this risk.

**Objective:** To determine the efficacy of pre-exposure prophylactic hydroxychloroquine for the prevention of SARS-CoV-2 infection and symptomatic coronavirus 19 disease (COVID-19) among HCWs at high occupational risk of SARS-CoV-2 exposure.

**Methods:** 130 HCWs in the New York University Langone Health System (NYULHS) who performed aerosol-generating procedures on patients with COVID-19 or provided bedside care for inpatients with COVID-19 or persons with suspected COVID-19 in an emergency department, for at least three shifts in a 7-day period, during the first 2020 COVID-19 wave in New York City were enrolled. Participants elected to take oral hydroxychloroquine, 600 mg on day 1 followed by 200 mg daily, or not take hydroxychloroquine for up to 90 days. Participants self-collected dried blood spots and completed digital questionnaires regarding COVID-19 symptoms, adverse events, and other COVID-19 medication use.

**Results:** Six participants (7.5%) seroconverted during the trial: four who took hydroxychloroquine (6.8%) and two who declined hydroxychloroquine (9.5%). All participants not taking hydroxychloroquine reported COVID-19 symptoms at seroconversion compared to one of four participants (25%) who took hydroxychloroquine. Adverse events occurred in eight participants (9.6%) on hydroxychloroquine and were mostly mild.

**Conclusions:** This study (ClinicalTrials.gov NCT04354870) did not demonstrate a statistically significant difference in SARS-CoV-2 seroconversion associated with hydroxychloroquine pre-exposure prophylaxis among HCWs at high risk of occupational SARS-CoV-2 exposure, although was underpowered and a high rate of hydroxychloroquine discontinuation was observed.

## Introduction

The coronavirus disease 19 (COVID-19) pandemic, caused by severe acute respiratory syndrome coronavirus 2 (SARS-CoV-2), has led to over 530 million cases and over 6.2 million deaths worldwide, including over 78 million cases and over 960,000 deaths in the United States.^1^ The mainstay of SARS-CoV-2 prevention throughout the pandemic focused on infection control measures, such as physical distancing, enhanced hand hygiene, and use of protective equipment. The development and implementation of effective COVID-19 vaccines significantly advanced COVID-19 prevention capacity, although breakthrough infections occur and vaccine rollout worldwide has been hindered by vaccine hesitancy and/or supply and distribution challenges. Multiple therapeutics exist for COVID-19, including immunomodulatory, antiviral, and monoclonal antibody agents; however, only one monoclonal antibody (tixagevimib/cilgavimab) is currently licensed as pre-exposure prophylaxis. Both the supply of tixagevimib/cilgavimab and the population eligible to receive it are limited. Additional effective pre-exposure prophylactic medications against SARS-CoV-2 would mitigate the risk of infection, especially if future SARS-CoV-2 variants capable of eluding vaccine-induced immunity and escaping neutralization by monoclonal antibodies become widespread. Effective pre-exposure prophylaxis would benefit individuals at high risk of exposure or severe disease in addition to those with decreased vaccine responsiveness.

Hydroxychloroquine is approved by the U.S. Food and Drug Administration for the treatment and prevention of malaria and the treatment of rheumatoid arthritis, systemic lupus erythematosus and chronic discoid lupus erythematosus. Interest in using hydroxychloroquine for SARS-CoV-2 infection developed after hydroxychloroquine and its relative, chloroquine, were demonstrated to inhibit SARS-CoV-2 *in vitro*.^2, 3^ Multiple mechanisms for the inhibition of SARS-CoV-2 by hydroxychloroquine have been proposed, including impairment of spike protein binding to cellular gangliosides, blockade of virus transport from early endosomes to lysosomes, and increased intraorganellar pH.^2, 4-7^ Additionally, hydroxychloroquine has immunomodulatory effects that could modify COVID-19 disease by dampening the hyperinflammatory state induced by SARS-CoV-2 infection.^7, 8^ The U.S. Food and Drug administration issued an emergency use authorization (EUA) for hydroxychloroquine on 28 March 2020, but revoked the EUA on 15 June 2020 based on lack of clinical benefit during treatment of patients hospitalized with COVID-19. The majority of studies of COVID-19 treatment with hydroxychloroquine, either alone or combined with azithromycin, have not demonstrated improvement in clinical outcomes.^9-19^

*In vitro* studies demonstrate that inhibition of SARS-CoV-2 by hydroxychloroquine was greater when the drug was added prior to viral infection compared to post-infection.^3^ This raised questions as to whether hydroxychloroquine pre-exposure prophylaxis could decrease SARS-CoV-2 infection. Clinical trials of hydroxychloroquine pre-exposure prophylaxis to date have not demonstrated a difference in SARS-CoV-2 infection.^20-22^ Here we present the results of a nonrandomized, open-label clinical trial [Clinicaltrials.gov NCT04354870] of hydroxychloroquine pre-exposure prophylaxis among health care workers (HCWs) at high risk of occupational SARS-CoV-2 exposure in the New York University Langone Health System (NYULHS) initiated during the first 2020 wave of the COVID-19 pandemic in New York City, prior to COVID-19 vaccine availability, to determine the efficacy of pre-exposure prophylactic hydroxychloroquine for the prevention of SARS-CoV-2 infection and symptomatic COVID-19.

## Methods

### Study Design

To evaluate whether hydroxychloroquine pre-exposure prophylaxis reduced the incidence of SARS-CoV-2, we conducted a nonrandomized, open-label clinical trial (ClinicalTrials.gov NCT04354870) among HCWs in the NYULHS. The primary outcome was seroconversion to SARS-CoV-2. Secondary outcomes included COVID-19 illness severity, incidence of symptomatic and asymptomatic SARS-CoV-2 infection, and hydroxychloroquine tolerability/adverse events. All study visits occurred remotely due to the ongoing COVID-19 pandemic. Following a combined screening and enrollment visit, study visits were conducted every 30 days (+/- 7 days) to complete four study visits. The study was reviewed and approved by the New York University Grossman School of Medicine (NYUSOM) institutional review board (IRB) prior to study initiation.

### Study Arm Assignments

Participants chose whether they were included in the hydroxychloroquine arm or no hydroxychloroquine arm of the study. Verbal assessment of whether an eligible participant elected to take or declined hydroxychloroquine was obtained at enrollment. Participants were allowed to switch between study arms by contacting the study team or indicating a change in hydroxychloroquine use via digital questionnaire. Primary and secondary outcome analyses were performed based on original study arm assignment using Independent Samples T-test with Welch’s correction (GraphPad Prism 9.2).

### Participants

This study recruited adults ages 18 years and older employed as HCWs in the NYULHS who provided informed consent for study participation and were involved in aerosol-generating procedures on confirmed COVID-19 patients using appropriate personal protective equipment (PPE), provided direct bedside care to confirmed COVID-19 patients using appropriate PPE for three or more shifts in a 7-day period, and/or provided direct care to suspected COVID-19 patients in the emergency department or another inpatient unit using appropriate PPE for three or more shifts in a 7-day period. Participants were excluded from the hydroxychloroquine arm if they had any of the following: diagnosed with COVID-19; known hypersensitivity to hydroxychloroquine or chloroquine; stage 4 or higher chronic kidney disease, retinal disease, congenital prolonged QTc interval syndrome, or torsade de pontes; or concomitant use of amiodarone, digoxin, flecainide, procainamide, or propafenone.

### Sample Size

An anticipated sample size of 350 participants was calculated to have 80% power to detect a 15% difference in seroconversion between the study arms assuming a two-sided type I error rate of 0.05 and a seroconversion rate of 35% in the no hydroxychloroquine arm. We did not know in advance how many study participants would decide to take hydroxychloroquine, but estimated two-thirds of participants would elect to take hydroxychloroquine.

### Informed Consent, Screening, and Demographics

Interested individuals were prescreened by telephone. Following prescreening, individuals who appeared eligible were emailed electronic consent forms using the Research Electronic Data Capture (REDCap) system. A study coordinator reviewed the electronic consent form with the participant by phone and answered any questions about the study during the screening/enrollment visit. Participants consented via electronic signature while in verbal communication with the study team, after which demographics, employment information, and medical history were collected to verify eligibility. Additional medication and symptom questionnaires were distributed to participants via REDCap.

### Hydroxychloroquine Dispensing

Prescriptions for oral hydroxychloroquine tablets (600 mg on day 1 followed by 200 mg daily thereafter) were provided to an NYU-based pharmacy which dispensed the study medication from a dedicated study supply for participants who opted to take hydroxychloroquine. Study medication was either picked up by participants in-person or mailed directly to participants. 30-day prescriptions were prescribed and filled in a similar manner at study day 30 and 60 for participants who remained in the hydroxychloroquine study arm.

### Digital Questionnaires

Individualized links to online REDCap questionnaires were emailed to participants at each study visit (screening/enrollment and days 30, 60, and 90 following enrollment). Links provided access to questionnaires regarding any potential adverse events, dried blood spot (DBS) card completion, medication use (hydroxychloroquine and concomitant medications), and potential COVID-19 symptoms. Questionnaire responses were connected to individual participants only by study-assigned identification numbers. Any participant reporting a potential adverse event was subsequently contacted by a study investigator to collect additional data.

### Dried Blood Spot Collection

DBS cards were self-collected by participants at enrollment, and days 30, 60, and 90 after enrollment. Participants were provided with four DBS collection kits and written instructions for collecting and returning DBS cards via designated drop-off collection boxes located at each participating NYULHS facility. Each collection kit contained: written collection instructions, alcohol pads, lancets, gauze, bandages, pre-labeled DBS cards and sealable foil bags containing desiccant, and envelopes for drop-off. In brief, participants were instructed to wash their hands, clean the tip of their finger with an alcohol pad, and deploy a contact-activated lancet to elicit a drop of blood. The first drop of blood was wiped away and subsequent drops were lightly dabbed onto the DBS card until all five spots on the card were filled, after which a bandage was applied to the finger. The DBS card was allowed to air dry at room temperature for 3 hours and then sealed in the foil bag containing desiccant, which was placed in a biosafety hazard-labeled envelope and deposited at one of the designated drop-off sites within 2 weeks after collection.

### Dried Blood Spot Elution

A 6 mm hole-puncher was used to remove dried blood spots from the card. Each punched out spot was placed in a well of a 24-well plate containing 300 µl phosphate-buffered saline (PBS) with 0.05% Tween 20 (PBS-T) and 0.08% sodium azide and shaken for 4 hours at room temperature or overnight at 4°C. Eluates were transferred to microcentrifuge tubes and centrifuged for 2 minutes at 10,500 g prior to use.

### Dried Blood Spot ELISA Validation

IgM and IgG antibodies to SARS-CoV-2 S1 protein in sera and DBS collected simultaneously from NYULHS employees with or without a known history of SARS-CoV-2 infection, who provided informed consent to participant in a separate NYUSOM IRB-approved protocol for healthy volunteers, were assessed using an in-house developed SARS-CoV-2 S1 ELISA. Dried blood spots eluates and sera were heat inactivated at 56°C for 1 hour prior to use.

### SARS-CoV-2 S1 ELISA

96-well ELISA plates were coated with 1 µg/ml SARS-CoV-2 S1 protein (Sino Biological Inc. 40591-V08H) diluted in PBS and incubated overnight at 4°C. Plates were washed four times with PBS-T and blocked with PBS-T containing 4% non-fat milk and 5% whey for 1 hour prior to adding sera or DBS eluates. For DBS ELISA validation and controls, sera samples were diluted to a 1:50 concentration while DBS eluates were not diluted. Plates were washed four times with PBS-T after a 2-hour incubation period at room temperature. Horseradish-peroxidase conjugated goat-anti human IgG and IgM (Southern BioTech 2040-05 and 2020-05) was added in 1:2000 and 1:1000 dilutions, respectively. After incubating for 1 hour plates were washed four times with PBS-T and developed with TMB Peroxidase Substrate 3,3′,5,5′-34029) for 5 minutes, after which the reaction was stopped with 1M sulfuric acid or 1N hydrochloric acid. The optical density (OD) was determined by measuring the absorbance at 450 nm on a Synergy 4 (BioTek) plate reader. Heat-inactivated pooled sera from hospitalized individuals with COVID-19 served as positive controls and sera collected from healthy volunteers prior to the COVID-19 pandemic served as negative controls. All control samples were obtained from research protocols approved by the NYUSOM IRB.

## Results

### Trial Participation

A total of 156 participants were screened for the trial, of which one hundred and thirty participants enrolled between 29 April 2020–15 May 2020. Enrollment was closed on 21 July 2020 due to lack of further interest from potential participants. Eighty-three participants elected to take hydroxychloroquine and 47 declined hydroxychloroquine (Figure 1). Participants who elected to take hydroxychloroquine were more likely to be male (p=0.02) and had a median older age (p=0.001) compared to participants who declined hydroxychloroquine (Table 1). No significant differences in race (p=0.91) or ethnicity (p=0.21) were reported among individuals who elected to take or declined hydroxychloroquine. Of the one hundred and thirty enrolled participants, thirty-nine (30%) formally left the study, of which twenty-three (17.7%) were participant-initiated and sixteen (12.3%) were withdrawn by study investigators. Only ten participants (12%) who originally elected to take hydroxychloroquine remained on the medication at day 90.

**Figure 1:**
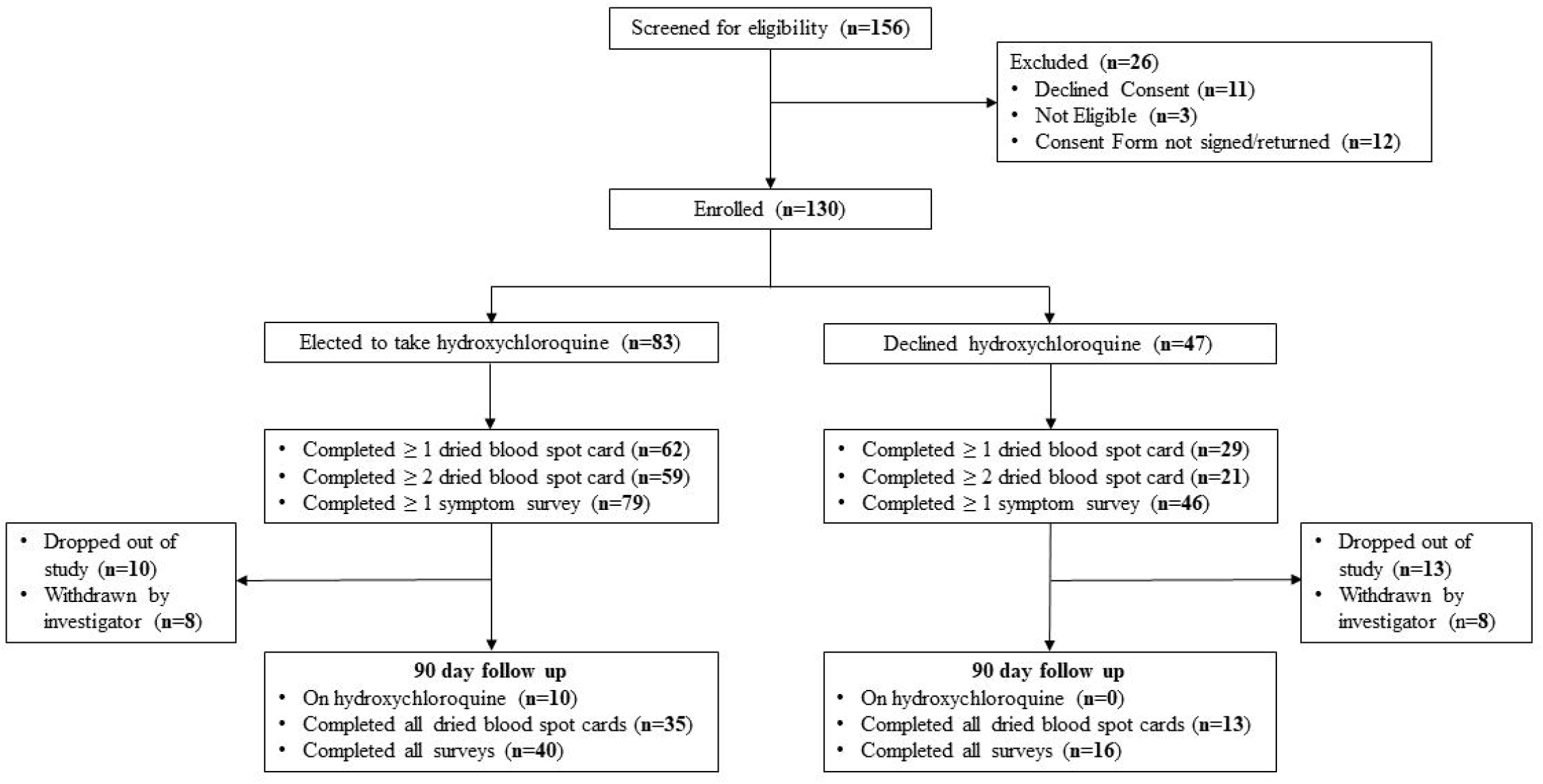
Flow Diagram of Participants in the Hydroxychloroquine Pre-Exposure Prophylaxis Study

**Table 1:** Enrolled Trial Participants According to Original Cohort Assignment by Age, Ethnicity, Gender, and Race

|  | Hydroxychloroquine<br>(n = 83) | No hydroxychloroquine<br>(n = 47) | All participants<br>(n = 130) | P value |
| --- | --- | --- | --- | --- |
| Number (%) by Gender |  |  |  |  |
| Female | 36 (43.4) | 30 (63.8) | 66 (50.8)<sup>a</sup> | 0.02<sup>b</sup> |
| Male | 46 (55.4) | 17 (36.2) | 63 (48.5)<sup>a</sup> |  |
| Other / Unknown | 1 (1.2) | 0 (0) | 1 (0.8)<sup>a</sup> |  |
| Median (IQR)<sup>c</sup> Age in Years | 39 (33, 50) | 32 (29, 38) | 36 (31, 47) | 0.001<sup>d</sup> |
| Number (%) by Race |  |  |  |  |
| American Indian/Alaska Native | 0 (0) | 0 (0) | 0 (0) | 0.91<sup>e</sup> |
| Asian | 13 (15.7) | 9 (19.1) | 22 (16.9) |  |
| Black / African American | 2 (2.4) | 2 (4.3) | 4 (3.1) |  |
| Native Hawaiian / Pacific Islander | 0 (0) | 0 (0) | 0 (0) |  |
| White | 58 (69.9) | 34 (72.3) | 92 (70.8) |  |
| Other / Unknown | 10 (12) | 2 (4.3) | 12 (9.2) |  |
| Number (%) by Ethnicity |  |  |  |  |
| Hispanic / Latinx | 5 (6) | 4 (8.5) | 9 (6.9) | 0.21<sup>e</sup> |
| Non-Hispanic | 59 (71.1) | 38 (80.9) | 97 (74.6) |  |
| Unknown | 19 (22.9) | 5 (10.6) | 24 (18.5) |  |

<sup>a</sup> Due to rounding, percentages do not add up precisely to 100%
<sup>b</sup> Independent Samples T-test
<sup>c</sup> Interquartile range
<sup>d</sup> Mann-Whitney U test
<sup>e</sup> Chi-squared test

### DBS SARS-CoV-2 S1 ELISA Validation

Eighty-two participating NYULHS employees consented to provide DBS samples from 27 March 2020–8 April 2020, forty with a previous diagnosis of SARS-CoV-2 infection (“experienced”) and forty-two with no known history of SARS-CoV-2 infection (“naive”). DBS ELISA data was normalized using a centering and scaling-type approach where the IgG and IgM optical densities (ODs) were scaled using the following formula: Scaled OD = (OD – average negative control) / (average positive control – average negative control) (Figure 2). Binary cut-off thresholds for scaled ODs between the two groups (SARS-CoV-2 “naive” and “experienced”) were identified at 0.61 for IgG and 0.49 for IgM. A significant correlation between the experienced and naive groups (p < 0.001) for both IgG and IgM was observed. Ten individuals from SARS-CoV-2 “naive” individuals tested positive for IgG and/or IgM, of which nine consented to have subsequent sera samples drawn which confirmed the presence of SARS-CoV-2 anti-spike antibodies (data not shown). On further questioning, four of these nine (44.4%) individuals recalled potential COVID-19 symptoms. The 23.8% seropositivity observed among “naïve” HCWs was consistent with 20% seropositivity observed in New York City in April 2020.^23^

**Figure 2:**
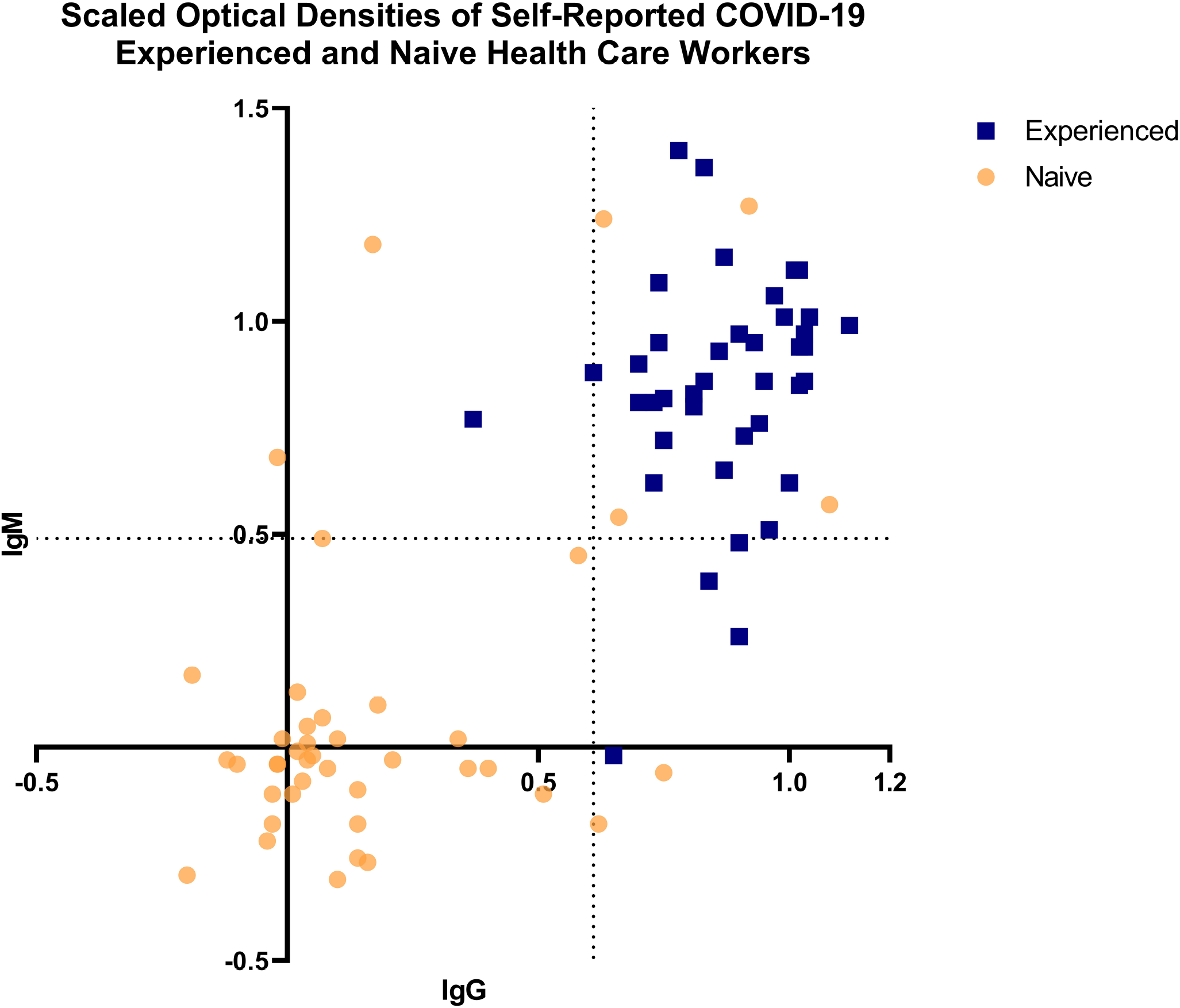
Dried Blood Spot ELISA Assay Validation — Scaled Optical Densities of Self-Reported COVID-19 Experienced and Naïve Health Care Workers These assays were performed during development of the DBS card antibody elution procedure and ELISA. Health care workers who reported a previous history of COVID-19 are shown as “experienced” blue squares, health care workers with no known history of COVID-19 are shown as “naive” yellow circles. Binary threshold cut-off values are indicated by the dotted lines.

### Seroconversion

Seroconversion was defined as a new positive IgM and/or IgG scaled antibody OD based on the binary cut-off criteria determined during ELISA validation. At least two DBS cards, the minimum required to assess for seroconversion, were returned by eighty participants (61.5%), fifty-nine of whom elected to take hydroxychloroquine (71.1%) and twenty-one of whom declined hydroxychloroquine (44.7%). Scaled ODs for IgM and IgG antibodies at all visits are shown in Figure 3. Six of the eighty participants (7.5%) who returned at least two DBS cards had seroconversion based on scaled ODs for either IgM and/or IgG anti-SARS-CoV-2 S1 antibodies during the trial, four of fifty-nine (6.8%) who elected to take hydroxychloroquine and two of twenty-one (9.5%) who declined hydroxychloroquine (p=0.71). Four participants (5%) had seroconversion of IgM antibodies only, three of fifty-nine who took hydroxychloroquine (5.1%) and one of twenty-one who declined hydroxychloroquine (4.8%). One participant in the no hydroxychloroquine arm (4.8%) developed seroconversion of both IgM and IgG and one participant on hydroxychloroquine (1.7%) developed only IgG seroconversion. Among the ninety-one study participants who returned at least one DBS card, five (5.5%) participants, all of which were in the hydroxychloroquine arm, had positive IgM or IgG ELISA results at study entry indicating previous SARS-CoV-2 infection.

**Figure 3:**
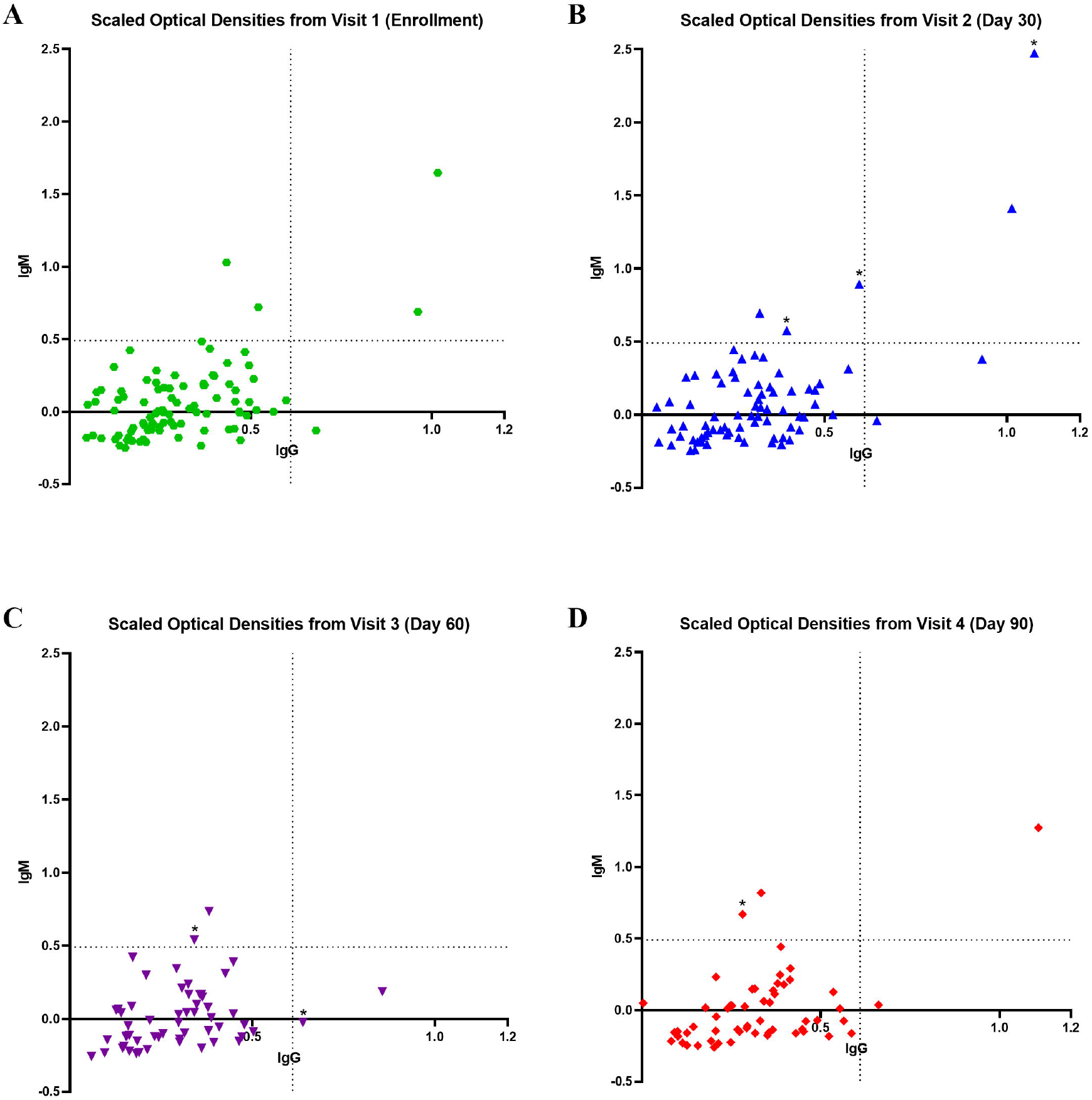
Scaled Optical Densities from Dried Blood Spots According to Visit Scaled optical densities from all returned dried blood spots from the enrollment visit are shown as green circles (A), from the day 30 visit as blue upward triangles (B), from the day 60 visit as purple downward triangles (C), and from the day 90 visit as red diamonds (D). Values representing new seroconversions are indicated with an asterisk above the value. Binary threshold cut-off values (established as in Figure 2) are indicated by the dotted lines.

### COVID-19 Symptom Assessment

At least one symptoms questionnaire was completed by one hundred and twenty-five participants (96.2%) and symptom questionnaires were completed at all four study time points by fifty-six participants (43.1%). Only one of four participants (25%) with seroconversion in the hydroxychloroquine arm reported potential COVID-19 symptoms (headache only) at the time of seroconversion. This compared to 100% (n=2, p=0.26) of participants with seroconversion in the no hydroxychloroquine arm who reported potential COVID-19 symptoms at the time of seroconversion: one who felt feverish and reported nausea/vomiting and sore throat, and the other with fever, chills, myalgias, headache, anosmia, cough, and shortness of breath. No participants were hospitalized or died during the study.

### Hydroxychloroquine Tolerability/Adverse Events

Adverse events were more common among participants originally assigned to the hydroxychloroquine arm [nine events among eight participants [9.6%, 95% CI: 3.2%-16.1%]) compared to none in the no hydroxychloroquine arm. The majority of adverse events were grade 1 (7 of 9, 77.8%) with one adverse event each with grade 2 (diarrhea) and grade 3 severity (rash) [table 2]. Grade 1 events included appetite loss, diarrhea, gastroesophageal reflux, gastrointestinal discomfort, hair loss, itching, and tinnitus. Among participants taking hydroxychloroquine, eight adverse events (88.9%) were classified as related to hydroxychloroquine use. Seven of the eight participants (87.5%) who experienced an adverse event discontinued hydroxychloroquine. No serious adverse events were reported.

**Table 2:** Adverse Events by Original Study Arm Assignment

| MedDRA Term | HCQ Group<br>n=83<br>n (%) | Control Group<br>n=47<br>n (%) |
| --- | --- | --- |
| **Appetite Loss** | 1 (1.2) | 0 (0) |
| *Grade I* | 1 (1.2) | 0 (0) |
| *Grade II* | 0 (0) | 0 (0) |
| *Grade III* | 0 (0) | 0 (0) |
| *Serious Adverse Events* | 0 (0) | 0 (0) |
| **Diarrhea** | 2 (2.4) | 0 (0) |
| *Grade I* | 1 (1.2) | 0 (0) |
| *Grade II* | 1 (1.2) | 0 (0) |
| *Grade III* | 0 (0) | 0 (0) |
| *Serious Adverse Events* | 0 (0) | 0 (0) |
| **Gastroesophageal Reflux** | 1 (1.2) | 0 (0) |
| *Grade I* | 1 (1.2) | 0 (0) |
| *Grade II* | 0 (0) | 0 (0) |
| *Grade III* | 0 (0) | 0 (0) |
| *Serious Adverse Events* | 0 (0) | 0 (0) |
| **Gastrointestinal discomfort** | 1 (1.2) | 0 (0) |
| *Grade I* | 1 (1.2) | 0 (0) |
| *Grade II* | 0 (0) | 0 (0) |
| *Grade III* | 0 (0) | 0 (0) |
| *Serious Adverse Events* | 0 (0) | 0 (0) |
| **Hair Loss** | 1 (1.2) | 0 (0) |
| *Grade I* | 1 (1.2) | 0 (0) |
| *Grade II* | 0 (0) | 0 (0) |
| *Grade III* | 0 (0) | 0 (0) |
| *Serious Adverse Events* | 0 (0) | 0 (0) |
| **Itching** | 1 (1.2) | 0 (0) |
| *Grade I* | 1 (1.2) | 0 (0) |
| *Grade II* | 0 (0) | 0 (0) |
| *Grade III* | 0 (0) | 0 (0) |
| *Serious Adverse Events* | 0 (0) | 0 (0) |
| **Rash** | 1 (1.2) | 0 (0) |
| *Grade I* | 0 (0) | 0 (0) |
| *Grade II* | 0 (0) | 0 (0) |
| *Grade III* | 1 (1.2) | 0 (0) |
| *Serious Adverse Events* | 0 (0) | 0 (0) |
| **Tinnitus** | 1 (1.2) | 0 (0) |
| *Grade I* | 1 (1.2) | 0 (0) |
| *Grade II* | 0 (0) | 0 (0) |
| *Grade III* | 0 (0) | 0 (0) |
| *Serious Adverse Events* | 0 (0) | 0 (0) |

## Discussion

Among health care workers at high risk of occupational SARS-CoV-2 exposure who elected to take oral hydroxychloroquine, 600 mg once followed by 200 mg daily, for up to 90 days, we did not observe a statistically significant decrease in SARS-CoV-2 seroconversion associated with hydroxychloroquine use. Study participants mostly identified as white and non-Hispanic, limiting the generalizability of the study results to races and ethnicities identified at higher risk for SARS-CoV-2 infection or disease.^24, 25^ However, these findings are consistent with those observed among other trials of hydroxychloroquine pre-exposure prophylaxis for HCWs and retrospective analyses of SARS-CoV-2 incidence among individuals taking hydroxychloroquine for other medical conditions.^20-22, 26-28^

Our methodology differed from previous studies in the dosing of hydroxychloroquine and the use of dried blood spots to assess for seroconversion with an in-house developed SARS-CoV-2 S1 ELISA. Dried blood spots were selected for use in this study due limitations in the availability of commercial serological SARS-CoV-2 testing at the time of study initiation and to facilitate self-collection of samples at home, thereby decreasing the risk of SARS-CoV-2 exposure by eliminating the need for travel to study visits. Although receipt of a COVID-19 vaccine could impact the interpretation of seroconversion of the S1 SARS-CoV-2 ELISA, this would not have affected the outcomes of this study as it was conducted in its entirety in 2020 prior to the availability of COVID-19 vaccines to the NYULHS HCW population.

The incidence of adverse events in the hydroxychloroquine arm in this study (9.6% of hydroxychloroquine users) was much lower than that reported (31%-45%) among other pre-exposure prophylaxis studies of HCWs.^20-22^ We do not anticipate the lower adverse event rate is attributable to the dosing selected, but could be impacted by our high rates of hydroxychloroquine discontinuation (88.0%) over the course of the study. We identify multiple factors that may have contributed to the high rate of hydroxychloroquine discontinuation in this study, including adverse events attributed to hydroxychloroquine, the U.S. Food and Drug administration revoking the EUA for hydroxychloroquine for COVID-19 treatment,^29^ and negative media coverage of hydroxychloroquine during the study period.

This study has several limitations. The study was underpowered to detect the effectiveness of hydroxychloroquine to reduce SARS-CoV-2 seroconversion. This study was initiated in April 2020, during the peak of the first wave of the pandemic in New York City.^30^ Initial estimates of anticipated seroconversion rates during the study period proved to be overestimates as the incidence of new COVID-19 cases significantly declined in New York City in May and June, 2020, and remained low for the remainder of the study period.^30^ Adoption of universal PPE use for all patient care encounters in the NYULHS during this period, including fit-tested N95 masks or equivalent, may have contributed to lower than expected seroconversion rates. Additionally, high rates of hydroxychloroquine discontinuation further decreased the ability of the study to detect differences in seroconversion between hydroxychloroquine pre-exposure prophylaxis users and non-users.

## Conclusions

Although our study was underpowered, our results suggest that pre-exposure prophylaxis with oral hydroxychloroquine, 600 mg once followed by 200 mg daily for up to 90 days, was safe but did not reduce seroconversion to SARS-CoV-2 among health care workers at high risk of occupational exposure to SARS-CoV-2 or reduce symptomatic presentation at the time of seroconversion. Further adequately-powered randomized controlled trials in areas with high SARS-CoV-2 circulation would be needed to adequately assess the effectiveness of off-label hydroxychloroquine for the prevention of SARS-CoV-2 infection.

## Data Availability

Individual level data from participants will not be shared to protect the privacy of the participating health care workers.

## Funding and Acknowledgements

The New York University Langone Health System provided funding for this study. This study was conducted using clinical support provided the NYU-H+H Clinical and Translational Science Institute (CTSI), which is supported by the National Center for Advancing Translational Sciences (NCATS), National Institutes of Health, through Grant Award Number UL1TR001445. We additionally wish to acknowledge technological support for electronic consenting for this study provided by the NYU Division of Advanced Research Technologies and the NYULH Office of Science and Research, as well as efforts provided by Marina Godina, Mahnoor Ali, Laura Frye, Tori Rodrick, and Spandan Pandya.

